# Emotional health concerns of oncology physicians in the United States: fallout during the COVID-19 pandemic

**DOI:** 10.1101/2020.06.11.20128702

**Authors:** Lauren Thomaier, Deanna Teoh, Patricia Jewett, Heather Beckwith, Helen Parsons, Jianling Yuan, Anne H. Blaes, Emil Lou, Jane Yuet Ching Hui, Rachel I. Vogel

**Affiliations:** University of Minnesota, Division of Gynecologic Oncology, Minneapolis, MN; University of Minnesota, Division of Hematology, Oncology and Transplantation, Minneapolis, MN; University of Minnesota, Division of Health Policy and Management, Minneapolis, MN; University of Minnesota, Department of Radiation Oncology, Minneapolis, MN; University of Minnesota, Department of Surgery, Minneapolis, MN

## Abstract

**Introduction:** Cancer care is significantly impacted by the Coronavirus Disease 2019 (COVID-19) pandemic. Our objective was to evaluate the effect of the pandemic on the emotional well-being of oncology providers across the United States and explore factors associated with anxiety and depression symptoms.

**Methods and Materials:** A cross-sectional survey was administered to United States cancer-care physicians recruited over a two-week period (3/27/2020 – 4/10/2020) using snowball-convenience sampling through social media. Symptoms of anxiety and depression were measured using the Patient Health Questionnaire (PHQ-4).

**Results:** Of 486 participants, 374 (77.0%) completed the PHQ-4: mean age 45.7±9.6 years; 63.2% female; all oncologic specialties were represented. The rates of anxiety and depression symptoms were 62.0% and 23.5%, respectively. Demographic factors associated with anxiety included female sex, younger age, and less time in clinical practice. Perception of inadequate PPE (68.6% vs. 57.4%, p=0.03) and practicing in a state with more COVID-19 cases (65.8% vs. 51.1%, p=0.01) were associated with anxiety symptoms. Factors significantly associated with both anxiety and depression included: degree to which COVID-19 has interfered with the ability to provide treatment to cancer patients and concern that patients will not receive the level of care needed for non-COVID-19 illness (all p-values <0.01).

**Conclusion:** The prevalence of anxiety and depression symptoms among oncology physicians in the United States during the COVID-19 pandemic is high. Our findings highlight factors associated with and sources of psychological distress to be addressed to protect the well-being of oncology physicians.

## Introduction

By early April 2020, the Coronavirus Disease 2019 (COVID-19) pandemic resulted in over 600,000 cases and over 24,500 deaths in the United States.^1^ Its burden on the healthcare system cannot be overstated. The pandemic has led to drastic changes in the delivery of health care: the creation of additional surge capacity for COVID-19 patients, and in oncology specifically, changes in work schedules and roles, rapid implementation of telehealth, delays of procedures, treatments, and ambulatory visits, suspension of clinical trials, changes in end-of-life care, and implementation of policies on the usage of personal protective equipment (PPE).^2^ Additionally, while cancer is considered a potential risk factor for COVID-19, it is not yet clear to what extent cancer increases the risk of being infected with COVID-19, or of developing complication in its disease course, though studies are starting to be published.^3-5^ These many changes and uncertainties in oncology practice affect individuals with cancer and their treating physicians. However, other than a few editorials or commentaries,^6,7^ no research to date has focused on the emotional well-being of physicians delivering cancer care in the United States.

While increased emotional strain and post-traumatic stress symptoms have been reported in COVID-19 frontline healthcare workers^8-10^, potential effects of the pandemic on other providers, such as oncologists, have received less attention. The drastic change in oncology practice due to COVID-19 and the uncertainty of its impact on the outcome of vulnerable cancer populations may cause significant stress among oncologists. We sought to evaluate the effect of the COVID-19 pandemic on the emotional health of oncology physicians across the United States and explore factors associated with anxiety and depression symptoms.

## Methods

We conducted a cross-sectional anonymous online survey among physicians who treat individuals with cancer in the United States. The study was reviewed and approved by the University of Minnesota Institutional Review Board. Eligibility criteria included: ≥18 years, ability to read/write in English, and being a physician (MD or DO) currently residing and providing cancer treatment to patients in the United States.

Individuals were recruited over a two-week period (March 27, 2020 – April 10, 2020) using snowball convenience sampling through social media (Twitter, Facebook, LinkedIn). Survey items included demographics, measures of clinical practice, concerns about COVID-19, effects of the pandemic on treatment decision-making and practice, and emotional well-being, using REDCap for data collection and storage.^11^ All questions were optional. We used the Patient Health Questionnaire (PHQ-4), a brief validated self-report screener to measure symptoms of depression and anxiety over the past two weeks.^12^ Potentially clinically relevant anxiety and depression symptoms were calculated using established cut-points of ≥3.^13,14^ The number of COVID-19 cases in each state was ascertained from the Centers of Disease Control and Prevention (CDC) as of April 3, 2020, the half-way point during the study recruitment period.^1^

Participant characteristics and responses were summarized using descriptive statistics, reporting means, standard deviations, and absolute and relative frequencies. We explored the associations of demographic, clinical practice, and COVID-19 related factors with self-reported depression and anxiety by comparing those with and without symptoms using Chi-squared tests and Fisher’ s Exact tests as appropriate for categorical variables, and t-tests assuming unequal variances for continuous variables. P-values <0.05 were considered statistically significant. Data were analyzed using SAS 9.4 (Cary, NC).

## Results

A total of 548 individuals started the survey; 62 were screened out as not eligible (32 not physicians who offer cancer treatments to patients, 30 not practicing in the United States). Of 486 eligible participants, 374 (77%) completed the PHQ-4 questions for inclusion in this analysis. Approximately two-thirds (63.2%) of respondents were female and the mean age was 45.7±9.6 years (Table 1). Just over half of respondents reported having minor children living with them, with females reporting this only slightly more than males (61.8% vs. 54.1%, p=0.34). Respondents practiced across 43 states with all oncologic specialties represented. Physicians reported treating a wide range of cancer types; the most common cancers treated were breast (58.5%), colon (38.8%), melanoma (32.1%), rectal (31.3%) and pancreatic (29.9%). More than half practiced in an academic setting, in a large city or its suburb, and at large or medium sized hospitals.

**Table 1.** Demographic and Clinical Practice Characteristics (N=374).

| **Characteristic** | **N** | **Mean±SD** |
| --- | --- | --- |
| **Age, years** | 337 | 45.7±9.6 |
| **Time in practice, years** | 357 | 12.9±10.1 |
|  | **N** | **%** |
| **Gender** |  |  |
| Male | 133 | 35.8 |
| Female | 235 | 63.2 |
| Non-binary gender identification | 4 | 1.1 |
| *Missing* | 2 |  |
| **Race/Ethnicity** |  |  |
| White, non-Hispanic | 272 | 75.1 |
| Asian Indian | 38 | 10.5 |
| Chinese | 16 | 4.4 |
| Hispanic | 12 | 3.3 |
| Other | 24 | 6.6 |
| *Missing* | 12 |  |
| **Children under 18 living with you** |  |  |
| No | 151 | 40.8 |
| Yes | 219 | 59.2 |
| *Missing* | 4 |  |
| **Medical Specialty** |  |  |
| Surgeon | 211 | 56.6 |
| Medical Oncology | 89 | 23.9 |
| Radiation Oncology | 54 | 14.5 |
| Other | 19 | 5.1 |
| *Missing* | 1 |  |
| **Practice at an academic institution** |  |  |
| No | 171 | 46.1 |
| Yes | 200 | 53.9 |
| *Missing* | 3 |  |
| **Hospital Size** |  |  |
| Small hospital (fewer than 100 beds) | 30 | 8.1 |
| Medium hospital (100-499 beds) | 160 | 43.0 |
| Large hospital (500 or more beds) | 170 | 45.7 |
| Ambulatory clinic only (no inpatients) | 12 | 3.2 |
| *Missing* | 2 |  |
| **Type of community (practice)** |  |  |
| Rural area | 18 | 4.9 |
| Small city or town | 81 | 21.8 |
| Suburb near a large city | 95 | 25.6 |
| Large city | 177 | 47.7 |
| *Missing* | 3 |  |

Most (74.8%) respondents were practicing in states with at least 1,000 confirmed COVID-19 cases at the time of the survey (Table 2). The majority (60.1%) reported being moderately or extremely concerned about getting COVID-19 and 20.3% considered themselves to be at high-risk for developing serious illness from COVID-19. Physicians reported COVID-19 had affected their ability to provide treatment to cancer patients to a moderately severe degree (66.8±24.6; severity range 0-100). A majority were moderately or extremely concerned about transmitting COVID-19 to a patient (65.9%) and the inability for their patients to receive the adequate level of care for a severe illness other than COVID-19 (70.5%).

**Table 2.** COVID-19 Exposure and Concerns (N=374).

| **Characteristic** | **N** | **%** |
| --- | --- | --- |
| **Number of COVID-19 cases in practicing state (as of 04/03/2020)** |  |  |
| 101-500 | 13 | 3.7 |
| 501-1000 | 77 | 21.6 |
| 1001-5000 | 133 | 37.4 |
| 5001 or more | 133 | 37.4 |
| *Missing (did not provide state where practice)* | 18 |  |
| **Do you consider yourself to be at “high risk” for severe illness from COVID-19?** |  |  |
| No | 257 | 68.7 |
| Yes | 76 | 20.3 |
| Unsure | 41 | 11.0 |
| **How concerned are you about getting COVID-19?** |  |  |
| Not at all concerned/Slightly concerned/Somewhat concerned | 149 | 40.0 |
| Moderately concerned/Extremely concerned | 224 | 60.0 |
| *Missing* | 1 |  |
| **How concerned are you about one of your family members getting COVID-19 from you?** |  |  |
| Not at all concerned/Slightly concerned/Somewhat concerned | 78 | 20.9 |
| Moderately concerned/Extremely concerned | 279 | 74.6 |
| Already happened | 2 | 0.5 |
| Not applicable | 15 | 4.0 |
| **How concerned are you about your patients getting COVID-19 from you?** |  |  |
| Not at all concerned/Slightly concerned/Somewhat concerned | 126 | 34.2 |
| Moderately concerned/Extremely concerned | 243 | 65.9 |
| *Missing* | 5 |  |
| **Adequate PPE for clinical practice** |  |  |
| No | 156 | 41.9 |
| Yes | 216 | 58.1 |
| *Missing* | 2 |  |
| **How concerned are you about your patients getting the level of healthcare they need if they become extremely ill from something other than COVID-19?** |  |  |
| Not at all concerned/Slightly concerned/Somewhat concerned | 110 | 29.5 |
| Moderately concerned/Extremely concerned | 263 | 70.5 |
| *Missing* | 1 |  |
|  | **N** | **Mean±SD** |
| **To what degree has COVID-19 interfered with your ability to provide treatment to active cancer patients? (0=no problem, 100=severe problem)** | 337 | 66.8±24.6 |

Almost two-thirds (62.0%) of oncology physicians in this study reported anxiety symptoms (Table 3). Demographic characteristics associated with anxiety included being female, younger age, and fewer years in clinical practice. Further, many items related to perceived COVID-19 risks were associated with anxiety, including practicing in a state with >1000 COVID-19 cases (65.58% vs. 51.1%, p=0.01) or having inadequate PPE access (68.6% vs. 57.4%, p=0.03). Almost one-quarter (23.5%) of respondents reported depression symptoms. While approximately a third who reported anxiety symptoms also reported depression symptoms (36.6%), almost all who reported depression symptoms were also anxious (96.6%). Asian-Indian physicians were more likely to report depression (36.8%) than non-Hispanic White (23.9%) or other race/ethnicity physicians (13.5%; p=0.04). Both anxiety and depression were associated with being moderately or extremely concerned about getting COVID-19, transmitting it to a family member or a patient, or inability for a patient to access an adequate level of care for a serious non-COVID-19 related illness (all p<0.01).

**Table 3.** Associations between demographic, clinical practice and COVID-19 variables and anxiety and depression symptoms (N=374).

|  | Anxiety* |  |  |  |  | Depression* |  |  |  |  |
| --- | --- | --- | --- | --- | --- | --- | --- | --- | --- | --- |
|  | No<br>N=142 (38.0%) |  | Yes<br>N=232 (62.0%) |  | p-value | No<br>N=286 (76.5%) |  | Yes<br>N=788 (23.5%) |  | p-value |
| Characteristic | N | Mean±SD | N | Mean±SD |  | N | Mean±SD | N | Mean±SD |  |
| Age, years | 125 | 47.6±10.7 | 212 | 44.6±8.7 | **0.008** | 256 | 46.1±9.8 | 81 | 44.7±8.8 | 0.23 |
| Time in practice, years | 136 | 15.0±11.6 | 221 | 11.5±8.8 | **0.003** | 272 | 13.3±10.4 | 85 | 11.5±9.0 | 0.11 |
| COVID-19 interfered with ability to provide treatment to active cancer patients?<br>(0=no problem, 100=severe problem) | 128 | 60.2±27.0 | 209 | 70.8±22.1 | **0.0002** | 259 | 65.3±25.0 | 78 | 71.5±22.7 | **0.04** |
|  | N | % | N | % | p-value | N | % | N | % | p-value |
| **Gender** |  |  |  |  | **<0.0001** |  |  |  |  | 0.32 |
| Male | 71 | 53.4 | 62 | 46.6 |  | 107 | 80.5 | 26 | 19.6 |  |
| Female | 67 | 28.5 | 168 | 71.5 |  | 174 | 74.0 | 61 | 26.0 |  |
| Non-binary gender identification | 2 | 50.0 | 2 | 50.0 |  | 3 | 75.0 | 1 | 25.0 |  |
| **Race** |  |  |  |  | 0.28 |  |  |  |  | **0.04** |
| White, non-Hispanic | 102 | 37.5 | 170 | 62.5 |  | 207 | 76.1 | 65 | 23.9 |  |
| Asian Indian | 10 | 26.3 | 28 | 73.7 |  | 24 | 63.2 | 14 | 36.8 |  |
| Other | 22 | 42.3 | 30 | 57.7 |  | 45 | 86.5 | 7 | 13.5 |  |
| **Children under 18 living with you** |  |  |  |  | 0.13 |  |  |  |  | 0.22 |
| No | 64 | 42.4 | 87 | 57.6 |  | 120 | 79.5 | 31 | 20.5 |  |
| Yes | 76 | 34.7 | 143 | 65.3 |  | 162 | 74.0 | 57 | 26.0 |  |
| **Medical Specialty** |  |  |  |  | 0.53 |  |  |  |  | 0.84 |
| Surgeon | 81 | 38.4 | 130 | 61.6 |  | 159 | 75.4 | 52 | 24.6 |  |
| Medical Oncology | 32 | 36.0 | 57 | 64.0 |  | 68 | 76.4 | 21 | 23.6 |  |
| Radiation Oncology | 24 | 44.4 | 30 | 55.6 |  | 44 | 81.5 | 10 | 18.5 |  |
| Other | 5 | 26.3 | 14 | 73.7 |  | 15 | 79.0 | 4 | 21.1 |  |
| **Hospital Size** |  |  |  |  | 0.19 |  |  |  |  | 0.71 |
| Small hospital (fewer than 100 beds) | 12 | 40.0 | 18 | 60.0 |  | 23 | 76.7 | 7 | 23.3 |  |
| Medium hospital (100-499 beds) | 68 | 42.5 | 92 | 57.5 |  | 126 | 78.8 | 34 | 21.3 |  |
| Large hospital (500 or more beds) | 58 | 34.1 | 112 | 65.9 |  | 125 | 73.5 | 45 | 26.5 |  |
| Ambulatory clinic only (no inpatients) | 2 | 16.7 | 10 | 83.3 |  | 10 | 83.3 | 2 | 16.7 |  |
| **Type of community (practice)** |  |  |  |  | 0.60 |  |  |  |  | 0.57 |
| Rural area | 5 | 27.8 | 13 | 72.2 |  | 13 | 72.2 | 5 | 27.8 |  |
| Small city or town | 33 | 40.7 | 48 | 59.3 |  | 64 | 79.0 | 17 | 21.0 |  |
| Suburb near a large city | 39 | 41.1 | 56 | 59.0 |  | 76 | 80.0 | 19 | 20.0 |  |
| Large city | 63 | 35.6 | 114 | 64.4 |  | 130 | 73.5 | 47 | 26.6 |  |
| **Practice at an academic institution** |  |  |  |  | 0.34 |  |  |  |  | 0.48 |
| No | 69 | 40.4 | 102 | 59.7 |  | 128 | 74.9 | 43 | 25.2 |  |
| Yes | 71 | 35.5 | 129 | 64.5 |  | 156 | 78.0 | 44 | 22.0 |  |
| **COVID-19 cases in practicing state\*\*** |  |  |  |  | **0.01** |  |  |  |  | 0.35 |
| 1000 or less | 44 | 48.9 | 46 | 51.1 |  | 72 | 80.0 | 18 | 20.0 |  |
| 1001 or more | 91 | 34.2 | 175 | 65.8 |  | 200 | 75.2 | 66 | 24.8 |  |
| **Do you consider yourself to be at “high risk” for severe illness from COVID-19?** |  |  |  |  | 0.29 |  |  |  |  | 0.58 |
| No | 104 | 40.5 | 153 | 59.5 |  | 200 | 77.8 | 57 | 22.2 |  |
| Yes | 26 | 34.2 | 50 | 65.8 |  | 57 | 75.0 | 19 | 25.0 |  |
| Unsure | 12 | 29.3 | 29 | 70.7 |  | 29 | 70.7 | 12 | 29.3 |  |
| **Concern about getting COVID-19** |  |  |  |  | **<0.0001** |  |  |  |  | **0.006** |
| Not at all concerned/Slightly concerned/Somewhat concerned | 88 | 59.1 | 61 | 40.9 |  | 125 | 83.9 | 24 | 16.1 |  |
| Moderately concerned/Extremely concerned | 53 | 23.7 | 171 | 76.3 |  | 160 | 71.4 | 64 | 28.6 |  |
| **Concern about family members getting COVID-19 from you** |  |  |  |  | **<0.0001** |  |  |  |  | **0.004** |
| Not at all concerned/Slightly concerned/Somewhat concerned | 50 | 64.1 | 28 | 35.9 |  | 70 | 89.7 | 8 | 10.3 |  |
| Moderately concerned/Extremely concerned | 89 | 31.9 | 190 | 68.1 |  | 208 | 74.6 | 71 | 25.5 |  |
| **Concern about your patients getting COVID-19 from you** |  |  |  |  | **0.002** |  |  |  |  | **0.007** |
| Not at all concerned/Slightly concerned/Somewhat concerned | 61 | 48.4 | 65 | 51.6 |  | 107 | 84.9 | 19 | 15.1 |  |
| Moderately concerned/Extremely concerned | 78 | 32.1 | 165 | 67.9 |  | 176 | 72.4 | 67 | 27.6 |  |
| **Adequate PPE for clinical practice** |  |  |  |  | **0.03** |  |  |  |  | 0.17 |
| No | 49 | 31.4 | 107 | 68.6 |  | 114 | 73.1 | 42 | 26.9 |  |
| Yes | 92 | 42.6 | 124 | 57.4 |  | 171 | 79.2 | 45 | 20.8 |  |
| **Concern about your patients getting the level of healthcare they need if they become extremely ill from something other than COVID-19** |  |  |  |  | **<0.0001** |  |  |  |  | **0.01** |
| Not at all concerned/Slightly concerned/Somewhat concerned | 59 | 53.6 | 51 | 46.4 |  | 94 | 85.5 | 16 | 14.6 |  |
| Moderately concerned/Extremely concerned | 83 | 31.6 | 180 | 68.4 |  | 192 | 73.0 | 71 | 27.0 |  |

*\*established cut-off for identifying potentially clinically relevant anxiety and/or depression using the PHQ-4*
*\*\* Centers for Disease Control, as of 04/03/2020*

## Discussion

Oncology physicians report significant anxiety and depression symptoms during the COVID-19 pandemic in the United States. Importantly, the perceived degree of interference with clinical practice along with personal concerns about COVID-19 were significantly associated with both anxiety and depression.

Previous studies from Wuhan, China have shown evidence of the high prevalence of distress, anxiety, and depression in COVID-19 “frontline” personnel, including nurses and physicians.^8,9^ Risk factors for emotional health symptoms included female sex, nursing profession, working on the “frontline,” and working in Wuhan, China.^9^ While it is not difficult to explain the pandemic’ s effects on the mental health of frontline healthcare workers, it is equally important to understand how the physician-based workforce and hospital care capabilities for other specialties have been affected. Oncology physicians have historically been more susceptible to burnout.^15^ and could be at even higher risk of distress during a crisis such as COVID-19. Wu et al. recently published a comparison of 190 oncology physicians and nurses who were either working in oncology or dispatched to the “frontline”.^10^ Those continuing to work in their usual capacity with uninfected cancer patients surprisingly had a greater frequency of burnout symptoms. Our findings of high anxiety among oncologists and the association of these symptoms with concerns about COVID-19 and patients getting adequate care for non-COVID-19 conditions are consistent with that study.

Strengths of this study include the national sample of oncologists treating a wide range of cancers at a time the pandemic was evolving in the United States. Limitations include its survey-based design, social media recruitment, and high proportions of surgeons and females; these respondents may not fully represent all oncologists practicing in the United States. We also chose to focus on physicians as they are heavily involved in treatment decision-making for oncology patients, however, we would expect similar results for advanced practice providers in oncology settings as well. These data may also under-report emotional symptoms due to social desirability bias. Finally, we do not know the rates of anxiety and depression among these oncologists prior to the pandemic and cannot infer these high rates were directly caused by the pandemic.

The prevalence of anxiety-related symptoms we observed among oncology physicians in the United States is alarming. These findings support a recent call to action to address burnout, prior to the pandemic, to protect the mental health of all oncologists in order to preserve their ability to deliver high quality and efficient care to cancer patients at a time of unprecedented uncertainty^16^. Further studies are needed to identify the sources of psychological distress and assess the efficacy of interventions for physicians during and after the COVID-19 pandemic. We have made tremendous strides in cancer care over the past few decades and to lose oncologists to burnout and its sequelae will be an extra insult to the already tragic situation of cancer and COVID-19.

## Data Availability

Data may be requested from the corresponding author.

